# A survey on the nationwide prevalence of asbestos-related lung cancer in Japan

**DOI:** 10.1101/2024.10.27.24316218

**Authors:** Mariko Niino, Taro Tomizuka, Yuka Nishina, Yuichi Ichinose, Wataru Gonoi, Takahiro Higashi

## Abstract

**BACKGROUND:** An accurate estimate of the nationwide prevalence of asbestos-related lung cancer (ARLC) is necessary to adequately operate a compensation subsidy program for patients with ARLC. Our study aimed to estimate the proportion of patients with ARLC among patients with primary lung cancer, and describe the characteristics and distribution of ARLC.

**METHODS:** All facilities that treated patients diagnosed with lung cancer in 2016 were requested to submit computed tomography images of 10 patients randomly selected from the national databases of hospital-based cancer registries. ARLC was defined as pleural plaques (PPs) extending over one-quarter of the inner lateral chest wall or existing PPs accompanied by obvious lung fibrosis. We estimated the proportion and distribution of ARLC among primary lung cancer cases and compared the characteristics of ARLC with those of primary lung cancer.

**RESULTS:** Of 772 facilities that treated at least one patient with lung cancer, 370 provided 3,565 sets of CT images. Of these, 216 (6.1%) had PPs, and 86 (2.4%) met the compensation criteria. After sample weighting, 2.0% of all primary lung cancers were classified as ARLC in Japan. A higher percentage of patients with ARLC were male (94.2% vs. 68.6%; *P* < .01) and had more advanced-stage disease (stage III: 22.1% vs. 16.0%; stage IV: 44.2% vs. 39.8%; *P* =.05) than other primary lung cancers. A majority (53.5%) of patients with ARLC were diagnosed at designated cancer hospitals. The proportion of squamous cell carcinoma was higher in ARLC than in those with primary lung cancer (25.6% vs. 18.6%; *P* < .01).

**CONCLUSION:** The estimated number of patients with ARLC was larger than expected from the number of applicants in the compensation system for asbestos-related health damages (AHDRS). Consequently, countermeasures are required to accurately identify eligible compensation recipients.

**Highlight:**

- The estimated proportion of patients with ARLC among patients with lung cancer was 2%.
- The proportion of squamous cell carcinoma was higher in ARLC than in primary lung cancer.
- A higher number of ARLC cases than expected from the official reports of AHDRS compensation recipients in Japan was found. More accurate AHDR reports are needed

## Introduction

Asbestos is a well-known carcinogen classified as group 1 (carcinogenic) by the International Agency for Research on Cancer.(1) However, asbestos-related malignancy, either lung cancer or mesothelioma, is insidious because the time from asbestos exposure to the incidence of malignancy can span several decades.(2, 3) A Japanese study reported that the period from the first exposure to the appearance of asbestos-related lung cancer (ARLC) ranged from 5 to 71 years, with a median of 47 years.(4) A Swedish cohort study reported a mean latency period of >44 years.(5) Although the range of estimated incubation periods varied across the survey, the incidence rate of asbestos-related malignancies is expected to increase continuously in the coming years.(6) This expectation is based on the observation that the peak amount of imported asbestos rose during the 1980s.(7)

Although mesothelioma strongly associated with asbestos is rare, lung cancer is relatively common and accounts for the highest number of cancer deaths in men and the second-highest number of deaths in women in Japan.(8) Because lung cancer has many known risk factors, such as smoking(9) and air pollution,(10) the frequency of lung cancers caused by asbestos is uncertain, with a massive difference across studies(11, 12) ranging from 4% to 12%. While the government has initiated a rescue program for ARLC to compensate for the detrimental health effects of industrial asbestos use, the number of applications is considered insufficient to cover all eligible individuals(12). The Asbestos Health Damage Relief System (AHDRS) of the Ministry of Environment provides financial aid to patients who are not covered by workers’ compensation programs.(13) The subcommittee of the Central Environment Council in Japan, which oversees the operation of AHDRS, has also raised the concern that the number of applications appears smaller than the actual number of patients with ARLC.(12) To properly operate ARLC compensation programs, ARLC prevalence must be accurately assessed.

An accurate estimate of the prevalence of ARLC in the Japanese population has not been obtained because the unique characteristics that distinguish ARLC from primary lung cancer have not been identified, therefore ARLC is often not diagnosed. Determining the nationwide prevalence of ARLC requires collecting medical images from many facilities, which further makes the survey challenging to conduct. Several previous studies have estimated the frequency of ARLC, but the study sites were limited to specific facilities for occupational health diseases.(7, 14, 15) Thus, in this study, we accurately estimated the prevalence of ARLC among patients with primary lung cancer nationwide and described the distinguishing characteristics of ARLC.

## Methods

### Asbestos-Related Lung Cancer Criteria

To identify patients with ARLC, we applied the criteria for CT images established by the Central Environment Council Asbestos Health Damage Assessment Subcommittee (HDAS) for the purpose of determining eligibility for compensation. Cases that met at least one of the following criteria were defined as ARLC: 1) extension of pleural plaques (PPs) to more than a quarter of the inner lateral chest wall; and 2) presence of PP accompanied by a clear shadow of pulmonary fibrosis indicating pneumoconiosis, with nodular or irregular opacity in both lung fields. The inner lateral chest wall line was defined as a curve that runs ventrally from the sternal border and dorsally to the origin of the ribs. For multiple plaques (including plaques observed in the mediastinal pleura in the same section), the total length of the plaques was measured.

The justification of the criteria is as follows: According to the Helsinki Consensus, these criteria are considered equivalent to a cumulative exposure of 25 fiber-years.(16) A twofold increased risk of lung cancer is associated with retained fiber levels of 5 million amphibole fibers (>1 μm) per gram of dry lung tissue. This lung fiber burden is approximately equal to 5,000–15,000 asbestos bodies, consisting of asbestos fibers coated by iron-containing protein and mucopolysaccharide per gram of dry lung tissue.(17) PP is caused by inhalation of fibrous silicate minerals, such as asbestos or erionite. Yusa et al.(18) reported that, among the cases in which >5000 asbestos bodies per gram of dry tissue were detected, 75% showed that the extent of PP was >25% of the inner chest wall.

### Subjects

The target population was patients with primary lung cancer diagnosed from January to December 2016. The cases were obtained from a hospital-based cancer registry (HBCR) database bearing the codes indicating invasive cancer (behavioral code 3) of the lung (C340-C349 by ICD-O-3 topography code) for patients who underwent initial treatment at the registering hospital.

### HBCRs and cancer care hospitals

The operation of an HBCR is mandated as a condition for designation as a cancer care hospital. These hospitals were designated by the Ministry of Health, Labor, and Welfare to specifically provide cancer care in their respective communities or local regions. Some non-designated hospitals that play central roles in their localities also maintain an HBCR. The designated and the some non-designated hospitals submit data to the National Cancer Center annually, the central database of which is estimated to cover approximately 70% of new cancer cases in Japan.(19) Personal identifiers in the HBCR are removed and replaced with identifying labels assigned solely for tracing. A catalog linking the identifying labels and personal identifiers are maintained by each medical facility.

We requested to use data from 772 medical facilities that submitted lung cancer cases in 2016 to the National Cancer Center. Among the lung cancer cases, we randomly selected 10 patients from each facility and requested the participating hospitals to obtain chest computed tomography (CT) images for these patients. If fewer than 10 lung cancer cases were treated at a facility in 2016, all registered lung cancer cases were requested for enrollment in the survey. We collected CT data taken nearest the date of diagnosis of lung cancer into an electronic file.

### Evaluation of CT Images

CT images were evaluated in two phases. In the first phase, 12 pulmonologists, and 12 radiologists identified suspected ARLC cases. These first-phase readers checked for PPs. If present, the readers documented the extension of PPs, presence of calcification, anatomical location of the PPs, and presence of pulmonary fibrosis. Two readers independently analyzed every CT image. If the readings of the two readers differed, a third reader analyzed the relevant CT images. In the second phase, five radiologists who specialized in ARLC analyzed the CT images. They all had experience in serving on the approval board of ARHDS as a member of HDAS. The five radiologists reviewed cases suspected with meet ALRC criteria by at least one reader during the first phase. Difficult cases that the experts did not want to judge alone were further discussed at a review meeting. We prioritized the experts’ opinions on the images that had judgment disagreements between the first and second phases.

### Statistical Analyses

The percentages of patients with PPs and of those that satisfied the ARLC criteria among all sampled patients with lung cancer were calculated. To estimate the proportion of ARLC among patients with primary lung cancer, the percentages of patients with ARLC were weighted against the total number of patients with lung cancer for each enrolled facility. The weights in each facility were calculated by dividing the number of registered patients with primary lung cancer by the number of samples from each facility.

To explore the characteristics of patients with ARLC, we compared the ARLC group with the non-ARLC group (other types of lung cancer) with respect to the following parameters: sex, age, geographic region, diagnostic facility type, primary lung cancer site, histological type, and cancer stage. We performed the chi-square tests to compare proportions and performed Student’s t-test to compare means between two groups. All tests were two-sided, and a *P* value < .05 was considered statistically significant. To determine the distribution of patients with ARLC according to medical facility type, we categorized the facility where the patients were diagnosed with lung cancer as a designated cancer hospital, laborers’ hospital, or another type of hospital by reviewing the subject’s HBCR data and then compared the proportions. In addition, we compared differences in CT image readings between generalists and asbestosis specialists to gain insight on the difficulty of diagnosis.

All statistical analyses were performed using Stata15 (StataCorp LLC, College Station, TX, USA). This research was approved by the Institutional Review Board of the National Cancer Center, Japan (2018-193). We conducted the survey from October 2018 to March 2020. During the two-year review period, we accessed the collected anonymized chest imaging data. Since the imaging data were anonymized, the authors did not have access to information that could identify individual participants during or after data collection. We facilitated the disclosure of research information and accommodated opt-outs.

## Results

### Estimated Proportion of ARLC in Primary Lung Cancer

Figure 1 shows the number of CT images eligible for analysis. For this survey, 370 facilities were enrolled, and CT serieses of 3,585 patients were collected. Image data from 20 patients were excluded because the files submitted were corrupted; thus, data on 3,565 patients were used for analysis. CT images from 203 patients were excluded because no suitable chest CT images were available for analysis and the necessary imaging data series for the assessments were not included. A final total 3,362 cases were available for analysis. Table 1 shows the characteristics of the selected patients compared with those of all patients with primary lung cancer in the 2016 HBCR. Table 1 shows the characteristics of the selected patients compared with those of all primary lung patients in the 2016 HBCR. (n = 86,804).

**Table 1.** Characteristics of patients with primary lung cancer in HBCR.

| Category |  | 2016 HBCR |  | Patients for analysis |  |
| --- | --- | --- | --- | --- | --- |
|  |  | (n = 86,804) | (%) | (n = 3,565) | (%) |
| Sex | Male | 58,492 | 67.4 | 2,476 | 69.5 |
|  | Female | 28,312 | 32.6 | 1,089 | 30.6 |
| Age at diagnosis, years | Under 39 | 477 | 0.6 | 13 | 0.4 |
|  | 40–49 | 2,049 | 2.4 | 69 | 1.9 |
|  | 50–59 | 6,460 | 7.4 | 224 | 6.3 |
|  | 60–69 | 25,918 | 29.9 | 955 | 26.8 |
|  | 70–79 | 33,643 | 38.8 | 1,308 | 36.7 |
|  | 80–89 | 16,729 | 19.3 | 879 | 24.7 |
|  | Over 90 | 1,528 | 1.8 | 117 | 3.3 |
| Histological type | Small-cell carcinoma | 7,326 | 8.4 | 327 | 9.2 |
|  | Squamous cell carcinoma | 16,701 | 19.2 | 668 | 18.7 |
|  | Adenocarcinoma | 47,547 | 54.8 | 1,758 | 49.3 |
|  | Other | 15,230 | 17.6 | 812 | 22.8 |
| Primary site of lung cancer | Bronchus | 3,207 | 3.7 | 183 | 5.1 |
|  | upper lobe | 43,609 | 50.2 | 1,747 | 49 |
|  | middle lobe | 4,724 | 5.4 | 168 | 4.7 |
|  | lower lobe | 33,512 | 38.6 | 1,360 | 38.2 |
|  | border | 56 | 0.1 | 4 | 0.1 |
|  | NOS | 1,696 | 2 | 103 | 2.9 |

HBCR, hospital-based cancer registry; NOS, not otherwise specified.

**Figure 1.**
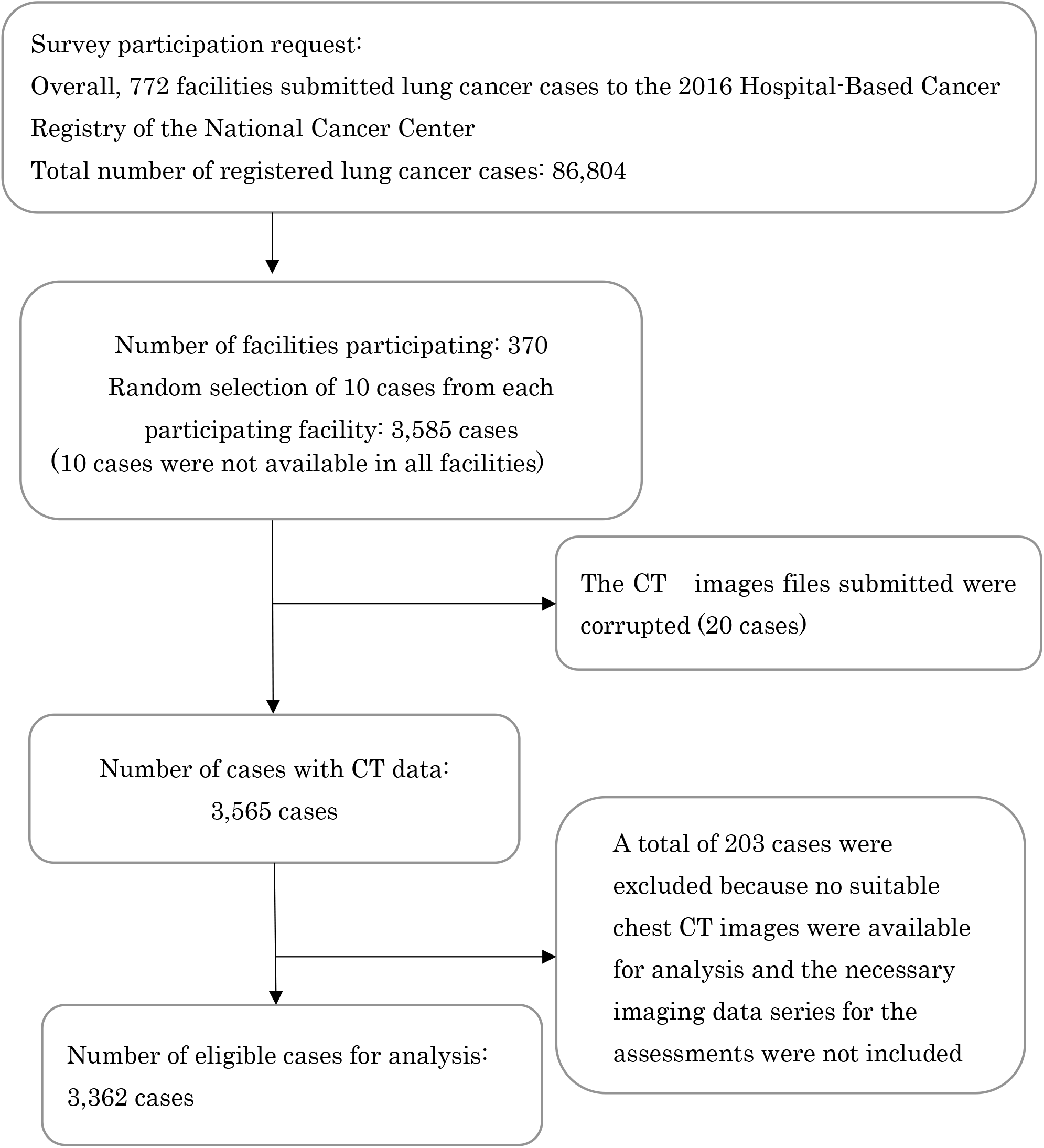
Flow chart of patient inclusion in the survey.

Of the 3,362 patients, 216 (6.4%) were diagnosed with PPs, and 86 (2.6%) met the criteria for ARLC. In 12.1% of cases, the presence of PPs could not be determined from the CT images. These cases were classified as not meeting the criteria for ARLC due to no clear fibrosis identified. PPs were not detected in 80.6% of patients with lung cancer (Figure 2-1). Among the 86 participants with PPs, 69 (80.2%) had more than a one-quarter spread of PPs, whereas 17 (19.8%) had PPs with obvious pulmonary fibrosis. After applying weightings, 2.0% of the patients with primary lung cancer registered in the 2016 HBCR were estimated to have ARLC (Figure 2-2).

**Figure 2.**
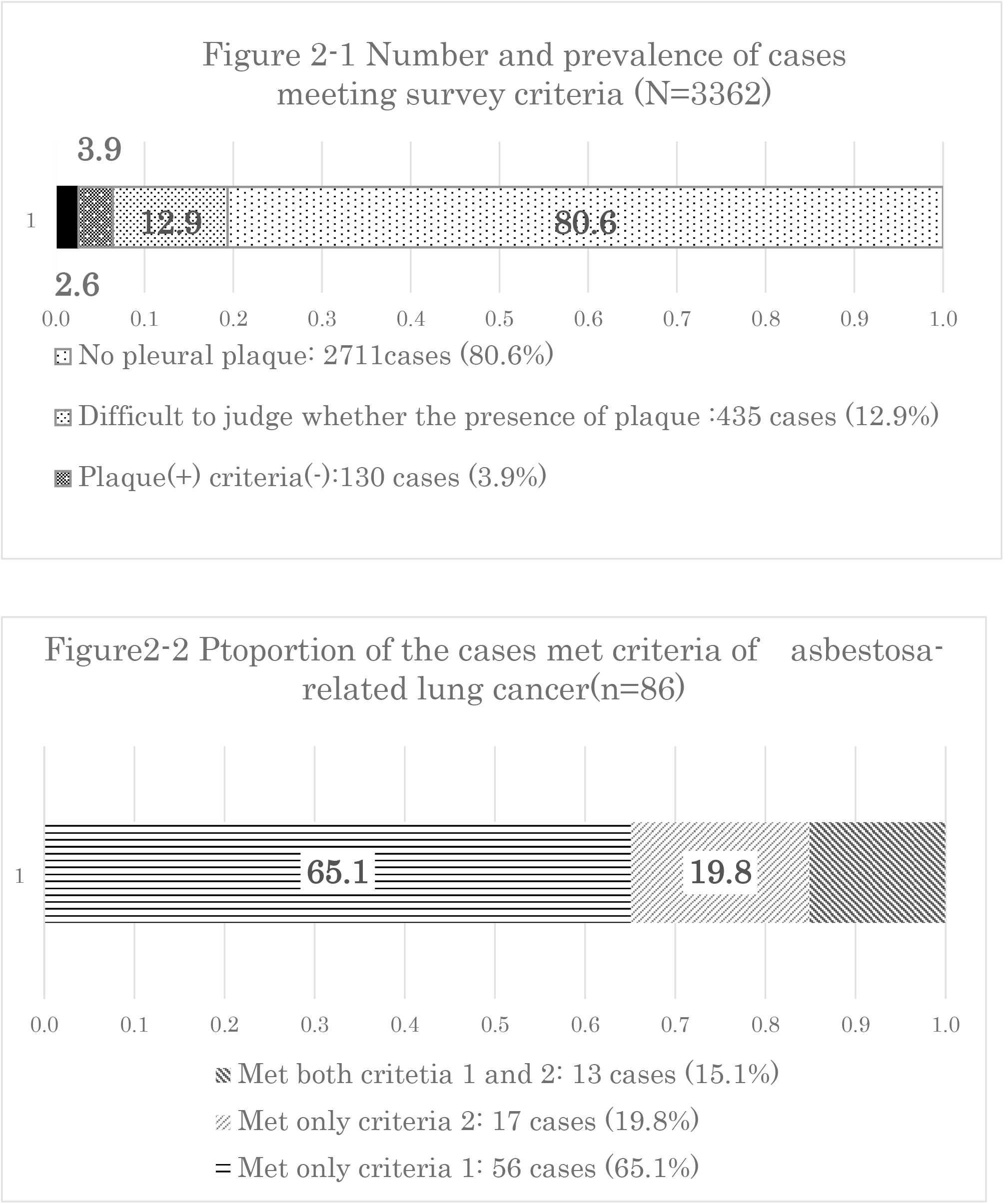
Number and prevalence of cases meeting the survey criteria.

### Differences in Criteria Judgment Between the Physicians and the Radiological Experts for ARLC

Radiologists in the second phase with experience in assessing ARHDS compensation recipients using the ARLC criteria, reviewed 503 cases; at least one reader suspected extension PPs or satisfaction with the ARLC criteria on CT images determined during the first reading phase, including 134 cases judged as meeting the ARLC criteria. The physicians and radiologists in first phase and radiological experts for ARLC differed in their assessments on 86 cases. Of these, 67 cases were rejected by the radiological experts, whereas 19 were rejected by the first phase physicians but judged by the experts as meeting the criteria for pulmonary fibrosis or PP extension. The main reason for rejection by the experts (41 cases) was misreading of PPs on CT by the physicians.

### Demographic and Histological Characteristics of Patients With ARLC

The characteristics of patients with and without ARLC are shown in Table 2. More men were in the ARLC group than in the non-ARLC group (94.2% vs. 68.6%; *P* < .01). The age at diagnosis tended to be higher for the patients with ARLC than for those with other types of lung cancer (71.7 vs. 68.2 years; *P* < .01). The proportion of squamous cells was significantly higher in the ARLC group than in the non-ARLC group (25.6% vs. 18.6%; *P* < .01) (Table 2).

**Table 2.**
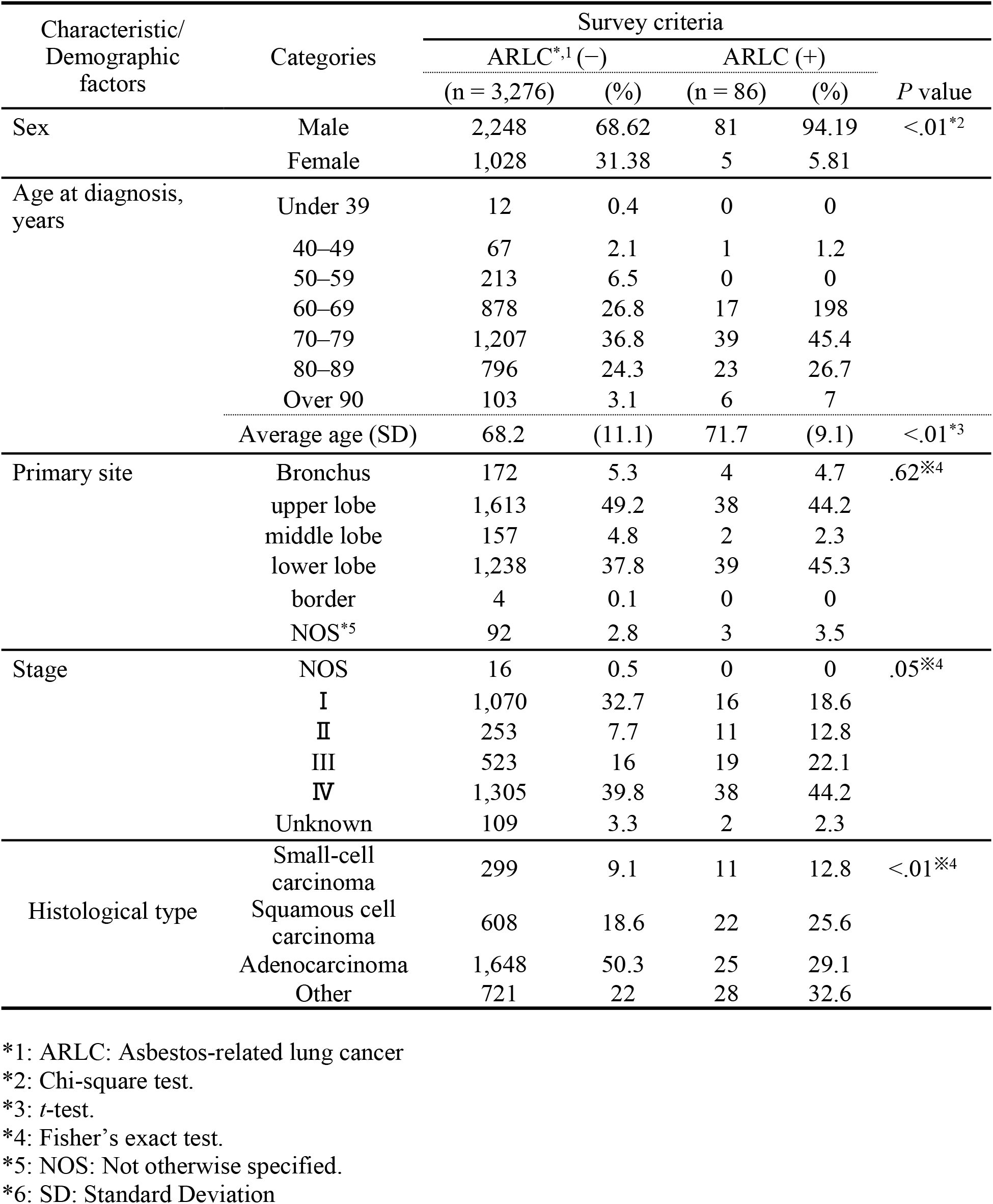
Comparison of the demographic and histological characteristics of patients with asbestos-related lung cancer and patients with other lung cancers.

### Distribution of ARLC Cases by Type of Medical Facility

The percentage of patients with ARLC among patients with lung cancer at the labor accident hospital (Rosai Hospital) was higher than those of other hospitals (8.2% vs. 2.4%; P < .01). However, 53.5% of the patients with ARLC were diagnosed at a designated cancer hospital, whereas 37.2% were diagnosed at other medical facilities.

## Discussion

Our study showed that 2% of patients with lung cancer met the criteria for ARLC. Based on the annual incidence of lung cancer of approximately 80,000 cases,(20) we expect 1600 patients to be diagnosed with ARLC annually, which is much larger than the total number of applications made to AHDRS and for workers’ compensation insurance. In 2016, 431 people were compensated by workers’ compensation and 134 by AHDRS.(21, 22) This finding might suggest that there are a significant number of patients who have been inaccurately diagnosed with ARLC. A lack of awareness among healthcare professionals regarding AHDRS for ARLC application might also contribute to discrepancies between the estimated numbers of our results and the actual count of compensation recipients. In addition, the estimated percentage of ARLCs highlights the number of potential patients who have not applied for asbestos compensation relief. Similarly, studies in other countries have reported that barriers to rescue systems for asbestos victims are related to underreporting.(23) Not only a lack of awareness of reportable conditions,(24) but also time constrains associated with completing multiple record requirements and involvement in government or legal hassles can contribute to underreporting by physicians(25) Moreover, physicians tend to underreport ARLC, especially if the patient is a smoker.(26)

The radiological imaging criteria above indicate that the patient had had cumulative asbestos exposure, increasing the risk of lung cancer twofold.(27) These criteria are useful because, although, there is a correlation between cumulative asbestos exposure and the risk of lung cancer,(28) accurately determining the level of exposure solely through exposure histories interviews is challenging due to its reliance on patient’s memories. In Japan, under the AHDRS, lung cancer is treated as asbestos-related cancer when the above criteria are met.

In previous studies in Japan that estimated the percentage of ARLC cases, the study sites were medical facilities specialized for work-related diseases; therefore, their estimates were expected to be higher than the proportions in a general population.(14) The present study included non-specialized medical facilities, which make our results more generalizable than those of previous studies. Furthermore, we found that a large proportion of patients with ARLC are diagnosed at regular designated cancer care hospitals, not at facilities specializing in work-related diseases. If we are to evaluate ARLC incidence appropriately, we should include non-specialized hospitals.

Notably, 17% (86/503 cases) of diagnoses differed between physicians in first phase and radiological experts for ARLC in second phase. A comparison of diagnoses between result of the first and second phases revealed that non-experts for ARLC physician more frequently made false-positive diagnoses. The discrepancies were mainly because of differences in determining the presence or absence of PPs. Several possible reasons for these discrepancies are inferred. First, general physicians have few opportunities to compare and discuss the interpretation of PPs on chest CT scans in clinical practice because PPs are not usually related to lung cancer treatments. Second, the pleural thickness and intercostal veins of pulmonary lesions caused by contact of the chest wall present characteristics similar to PPs; therefore, making them difficult to distinguish. In addition, PPs are sometimes difficult to distinguish from false-positive findings on radiological images. The survey results indicate that a certain number of cases are rejected during AHDRS accreditation by physicians who submit false-positive findings. Because the paperwork is substantial, a false-positive finding would waste the time of both physicians and patients. Further continuing education on the accurate diagnosis of PPs before submitting applications to the AHDRS is needed. Additionally, potential strategies include promoting application submissions even with false positives to minimize omissions and exploring the development of automated detection systems.

We found a higher proportion of squamous cell carcinomas in ARLC than in non-ARLC(35.6% vs. 18.6%). Our results showed that the proportion of squamous cell carcinomas among ARLC cases was relatively higher, which might have been influenced by the higher proportion of smokers in the asbestos-related disease cohort than in the general population since smoking synergistically increases the risk of asbestos-related lung disease, and the said cohort has a high risk of developing squamous cell carcinoma.(29) A comparison of cancer stages at the time of diagnosis showed that ARLC tended to be more advanced on diagnosis than other lung cancers (stage III: 22.1% vs. 16.0%; stage IV: 44.2% vs. 39.8%). Generally, the prognosis is poor in ARLC because the cancer develops in older patients because of the long incubation period and low lung function due to lung fibrosis and emphysema.

### Limitations

Our study has several limitations. First, we only analyzed CT images from patients who consented to the use of their data. The response rate was 55%, which is reasonable since we conducted a national survey. However, the number may not be representative of the population with lung cancer in Japan. In spite of this, we included as wide a variety of hospitals as possible and implemented a random sampling design. Second, the incidence of ARLC may be changing chronologically. We surveyed patients diagnosed in 2016, which indicates that repeated studies are warranted to show any potential trend over time. Third, we only collected a limited amount of background information on the patients. We placed a higher priority on recruiting hospitals to estimate the prevalence of ARLC as accurately as possible and did not collect information exhaustively to avoid burdening the hospitals. However, this limited our ability to perform more detailed analyses, such as assessing smoking status. Finally, to simplify the data collection process, we did not collect simple chest radiography and judged fibrosis based on the CT images, which may have overestimated the incidence of the second criteria because CT is much more sensitive than chest X-ray imaging in detecting fibrosis. However, the potential overestimation would not change our results much because >80% of the patients met the criteria of having widespread PPs.

## Data Availability

Although the data used in this study are anonymized when obtained from facilities, we have not obtained consent from the participants to share the data with third parties or to make the data public. Therefore, the use of the data is restricted.

## Abbreviations list

ARLC: asbestos-related lung cancer
AHDRS: Asbestos Health Damage Relief System
CT: computed tomography
AHDAS: Asbestos Health Damage Assessment Subcommittee
HRCR: Hospital-Based Cancer Registry
PPs: pleural plaques

